# Multi-Year Comparison of VITEK® MS performance for identification of rarely encountered pathogenic gram-negative bacilli (GNBs) in a large integrated Canadian healthcare region

**DOI:** 10.1101/2024.08.22.24312438

**Authors:** D.L. Church, T Griener, D Gregson

**Affiliations:** Department of Pathology & Laboratory Medicine, Cummings School of Medicine, University of Calgary, 3330 Hospital Dr. NW, Calgary, Alta. T2N 4B1; Department of Medicine, Cummings School of Medicine, University of Calgary, 3330 Hospital Dr. NW, Calgary, Alta. T2N 4B1; Alberta Precision Laboratories, Research Rd. NW, Calgary, Alta. T2L 2K8, 9-3535, Canada

**Keywords:** identification, Gram-negative bacilli, VITEK MS, proteomics

## Abstract

**Background:** This multi-year study (2014–19) compared identification of rare and unusual GNB by MALDI-TOF MS (VITEK® MS, bioMérieux, Laval Que.) to 16S rRNA gene sequencing (16S) according to our laboratories routine workflow; 16S is done if initial MALDI-TOF MS results were discordant, wrong or absent.

**Materials and Methods:** GNB isolates were first analyzed by standard phenotypic methods and MALDI-TOF MS using direct deposit with full formic acid extraction; proteomics was repeated if no result occurred. Medically approved 16S analyses were done using fast protocols. Isolate sequences were analyzed using IDNS3 bacterial database (SmartGene™, Lausanne, Switzerland).

**Results:** 329 GNB isolates were recovered from 304 specimens; >1 isolate was recovered from 19(6%). 250(76%) NFGNBs, 62(19%) fGNBs, and 17(5%) CAMPB were mainly recovered from blood cultures (31.6%) and lower respiratory specimens (43%) (one-half were isolated from cystic fibrosis patients). Accurate genus vs. species identities were obtained for 67.2%/26% NFGNBs, 74.2%/53.2% fGNBs, and 22% CAMPB (with no discrepant species), respectively. Wrong or no results were obtained for 82(32.8%) NFGNB, 17(27.4%) fGNB, and 13(72.2%) CAMPB. Absent or misidentifications occurred for NFGNBs (33%), fGNBs (26%) and CAMPB (89%) due to absence of species in the instrument’s database. VITEK MS performance remained stable for NFGNBs and fGNBs but improved for CAMPB but with the addition of *Campylobacter rectus* and *Campylobacter curvus* to the database.

**Conclusions:** VITEK® MS databases need to be continually updated to include an increasing number of rare and unusual GNBs causing invasive human infections. 16S remains important for identification of GNBs where proteomics fails.

## Introduction

Gram-negative bacilli are commonly encountered pathogens in clinical microbiology laboratories. Non-fermenting gram-negative bacteria (NFGNB) are largely environmental opportunists causing severe infections in immunocompromised individuals(1–5). Pathogenic colonization in cystic fibrosis patients, especially with *Pseudomonas aeruginosa*, results in decline in pulmonary function and potential morbidity and mortality(6–8). Human gut microbiome includes enteric gram-negative bacteria (GNBs) which cause urinary tract or intra-abdominal infections with associated bloodstream infection(1, 9, 10). HACEK group GNBs (*Haemophilus*, *Aggregatibacter*, *Cardiobacterium*, *Eikenella* and *Kingella*) are rare causes of infective endocarditis but or fastidious GNBs (fGNBs) cause different infections(11, 12). Campylobacterales (CAMPB) cause gastrointestinal infections including peptic ulcer disease (i.e., *Helicobacter pylori*) but rarely cause or disease(13–16). Effective therapy requires accurate identification of pathogenic GNBs due to their unique antibiotic susceptibility profiles, and inherent or developing multi-drug resistance(9, 17–19).

Definitive identification of GNBs is sub-optimal using phenotypic methods because of low biochemical reactivity. Accuracy and timeliness of GNB identification along with patient outcomes has improved with use of matrix-assisted laser desorption ionization-time of flight mass spectrometry (MALDI-TOF MS)(20, 21). MALDI-TOF MS databases currently lack spectral profiles for some unusual and rarely encountered pathogenic GNBs resulting in isolate misidentification or lack of identification. Limited studies compared VITEK® MS (MALDI-TOF MS) (bioMérieux, Laval, Que.) to 16S for identification of unusual or rarely encountered pathogenic GNBs(22–25). Prior reports of MALDI-TOF MS performance used GNB isolates that had prior 16S analysis to include organisms available in proteomics databases(22–25). We verified performance of our laboratories workflow by comparing VITEK MS results since implementation in 2014 to secondary 16S analysis of isolates that proteomics misidentified. We encounter a highly number of unusual or rarely invasive GNB compared to smaller hospital laboratories in our regional network due to consolidated regional nature of our large referral facility.

## Materials and Methods

### 1.1 Patients and clinical specimens

Patients who had ≥1 GNB isolate from blood or non-blood invasive clinical specimens were enrolled over a multi-year period (2014–2019) from Calgary and South Zones, Alberta Health Services (AHS) following clinical review by a medical microbiologist and Infectious Diseases specialist. Adults had two sets of blood cultures (i.e., each consisting of an aerobic/anaerobic bottle) according to Calgary Zone regional protocol. Non-blood specimens (abscesses, tissues, sterile fluids) were collected operatively or by interventional radiology under ultrasound guidance. Non-blood specimens were collected using standard collection devices and protocols to ensure recovery of GNB. Clinical specimens were transported for immediate processing to laboratory within 2h after collection.

### 1.2 Laboratory setting

Our large regional laboratory in Calgary Zone, Alberta Health Service does clinical microbiology testing for Calgary and surrounding hamlets and towns in Southern Alberta representing an urban and rural population of ∼2.6 million people including adult tertiary hospitals, a tertiary pediatric hospital, several rural facilities, all ambulatory practices, and long-term care centers.

### 1.3 Laboratory analyses

Clinical specimens were initially analyzed using standard phenotypic methods including a microscopic examination and aerobic and anaerobic culture methods (except for sputa). Blood and sterile fluids cultured using an aerobic BACT/ALERT FA plus and FN plus bottle pair incubated for up to five days in a BacT/Alert instrument (bioMérieux, Laval, Quebec).

Subculture of positive blood cultures inoculated blood agar (BA), chocolate agar (CHOC), MacConkey agar (MAC) and Brucella blood agar (BBA) (Dalynn, Calgary) incubated for 5d at 35°C. Subculture of sputa and or deep lung samples [i.e. bronchoalveolar lavages (BALs), bronchial washes (BWs)] inoculated BA, CHOC and MAC incubated under aerobic conditions at 35°C for 4d. Tissue and deep abscess specimens were inoculated to BA, CHOC and BBA; BA and CHOC plates incubated in 5% C0_2_, MAC was incubated in 0_2_, and BBA incubated in an Anoxomat™ anaerobic jar system (Fisher Scientific, Mississauga, Ont.) for 4d. GNB isolates were confirmed as enteric, non-fermenting, fastidious or microaerophilic by standard phenotypic procedures (i.e., growth on specific media, atmospheric growth conditions, colony morphology, Gram stain reaction/morphology and rapid biochemical tests). GNB isolates were subsequently analyzed according to our laboratory’s workflow; MALDI-TOF MS is first done with subsequent 16S rRNA gene sequencing when proteomics gives a low confidence or absent result. Direct deposit full formic acid extraction was done for MALDI-TOF MS (VITEK® MS, bioMérieux, Laval, Quebec) analysis according to manufacturer’s instructions as described in previously published guidelines(26). Repeat MALDI-TOF MS was done with same method if initial results gave low confidence (<99%) or no identification. Reported VITEK® MS results had a high confidence (i.e., ≥99.0% or log score ≥2) and agreed with phenotypic results. VITEK® MS V 2.0 (2012) database was used from 2014 to 2016, V3.0 from 2016 to 18, and V3.2 until 2019. A total of 11478 NFGNB and fastidious and 55 Campylobacterales isolates were identified with high confidence using phenotypic plus VITEK® MS analysis during the study period and were not enrolled. Unusual or rarely encountered GNBs were enrolled and had molecular analysis when proteomics gave a wrong identification or failed to provide a result. 16S rRNA gene sequencing was performed by fast PCR/cycle sequencing using fast MicroSEQ™ 500 16S DNA PCR kits and an ABI Prism 3500 XL sequencer (Applied Biosystems, ThermoFisher Scientific, Foster City, CA) as previously described(27). SmartGene’s Integrated Database Network System (IDNS3®) (Lausanne, Switzerland) bacterial database was used to determine closely related species (https://www.Smartgene.com). Isolate sequences were compared to a well characterized reference sequence and overall identity scores were 99.9% (0-2 mismatches). 16S rRNA gene sequencing identification to genus- or species-level used interpretive criteria outlined in Clinical and Laboratory Standards Institute (CLSI), Approved Guidelines MM-18 for targeted DNA sequencing analysis(28).

### 1.4 Data analysis

Data were entered into a Microsoft Excel spreadsheet (MS Office 2016) and analyzed according to standard descriptive methods. VITEK MS performance was calculated against current instrument databases (i.e., V 2.0, V3.0 or V3.2) ability to accurately identify GNBs at time of study. VITEK MS performance was compared to ‘gold standard’ method (i.e., 16S rRNA gene sequencing).

### 2. Ethics, guidelines, and consent to participate

This study was approved, and a waiver of consent was granted by Conjoint Health Ethics Research Board (CHREB), Alberta Health Services, and University of Calgary (REB15-0629). This study complied with all relevant guidelines and regulations.

## Results

### 2.1 Clinical specimens and isolates

Patients’ diagnosis included bloodstream infection, pneumonia, lung abscesses, cystic fibrosis, deep wound infections, deep organ and intra-abdominal abscesses, and deep skin and soft tissue infections including burns. 329 GNB isolates were recovered from 304 specimens; >1 isolate was recovered from 19/304 (6%) specimens. Isolates studied included 250(76%) NFGNBs, 62(19%) fGNBs, and 17(5%) CAMPB. This represents 2.3% of NFGNBs, 1.7% of fGNBs and 31% of CAMPB isolates identified during the study period. **Figure 1** outlines recovery of isolates from clinical specimens; most were recovered from blood cultures (96/304; 31.6%) and lower respiratory site/sources (130/304;43%) including bronchoalveolar lavages (BALs), bronchial wash (BWs), sputum and throat specimens from cystic fibrosis. One-half of clinical respiratory specimens were submitted from patients with cystic fibrosis (CF). Enrollment of isolates remained constant with an average of 59 (range = 41 to 89) isolates accrued each per year (Data not shown).

**Figure 1.**
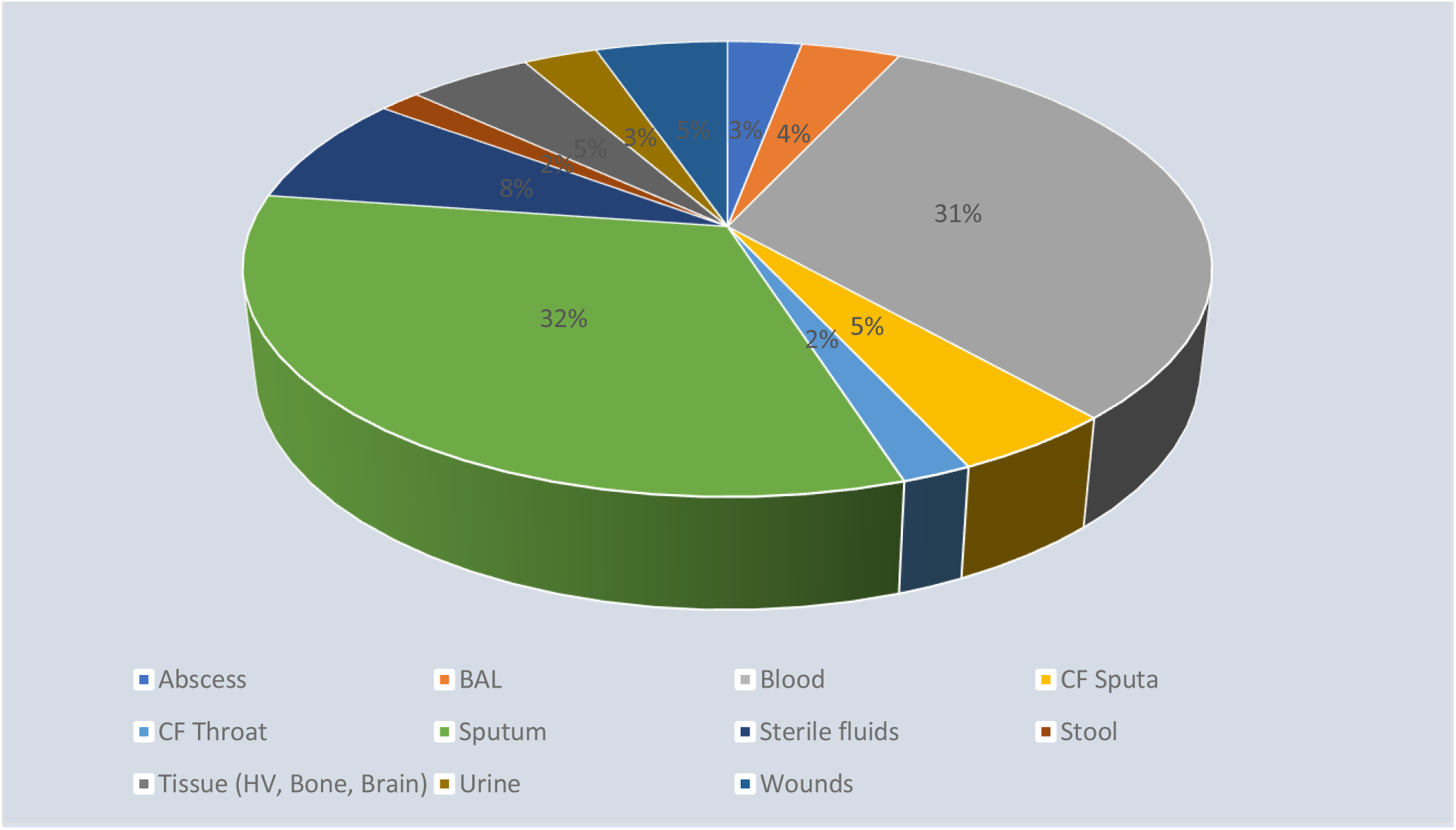
Recovery of rare and unusual Gram-nega琀椀ve bacilli from clinical specimens

### 2.2 Description of GNBs

**Table 1** describes 37 genera and 80 species of NFGNBs studied. Predominant genera were *Achromobacter* spp (n=28, 12%) and *Burkholderia* spp. (n=53, 22%) but a diverse spectrum of NFGNBs were represented. Predominant species included *A. xyloxidans* (n=25), *Bordetella* spp. (n=9) (*B. bronchiseptica*, *B. hinzii*, *B holmesii*, and *B. petrii*), *Chryseobacterium* spp. (n=11) (*C. aquifrigidense*, *C. hominis*, *C. indologenes*, and *C. profundimaris*), *Legionella* spp. (n=10) (*L. bozemanae*, *L. maceachernii*, and *L. pneumophila*), *Paracoccus* spp. (n=12) (*P. panacisoli* and *P. yeei*), *Pseudomonas* spp. (n=8) (*P. aeruginosa*, *P. alcaligenes*, *P. costantinii*, *P. moraviensis*, *P. nitroreducens* and *P. trivialis*), and *Sphingomonas* spp. (*S. paucimobilis* and *S. spiritivorum*) (**Table 1**). Accuracy of VITEK® MS NFGNB identification remained stable due to limited addition of new NFGNB spectral database profiles **Table 1**.

**Table 1.**
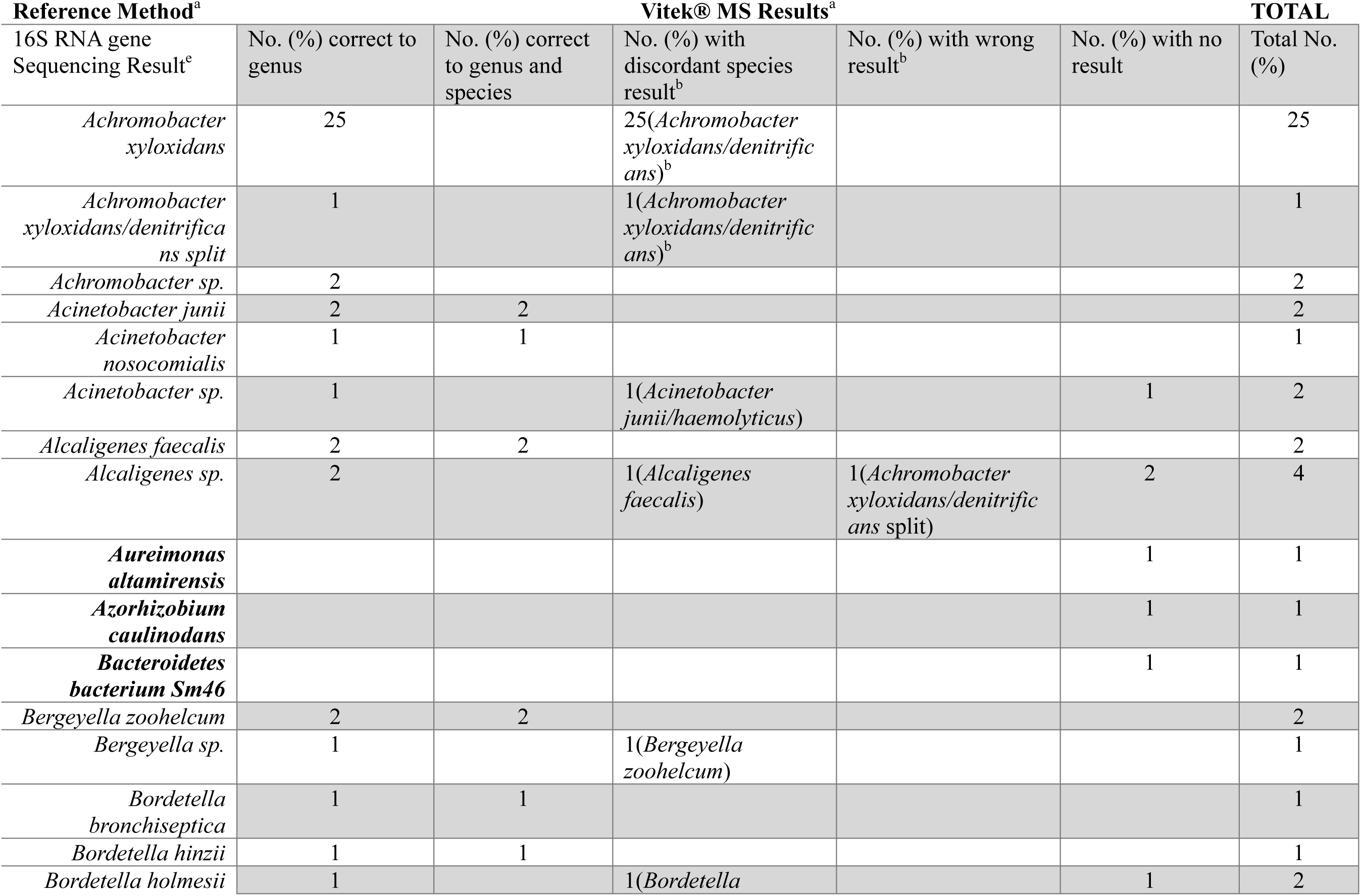
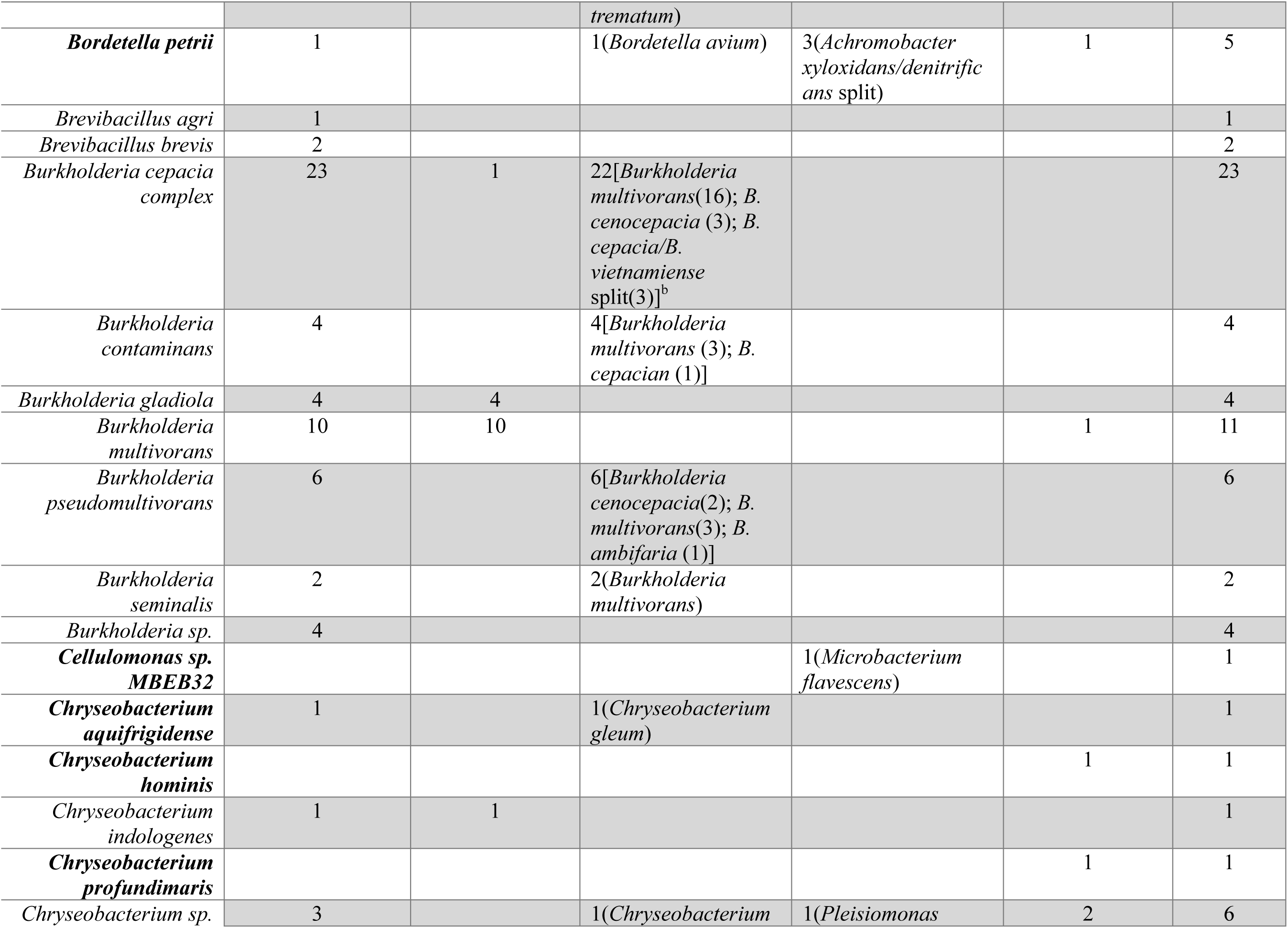
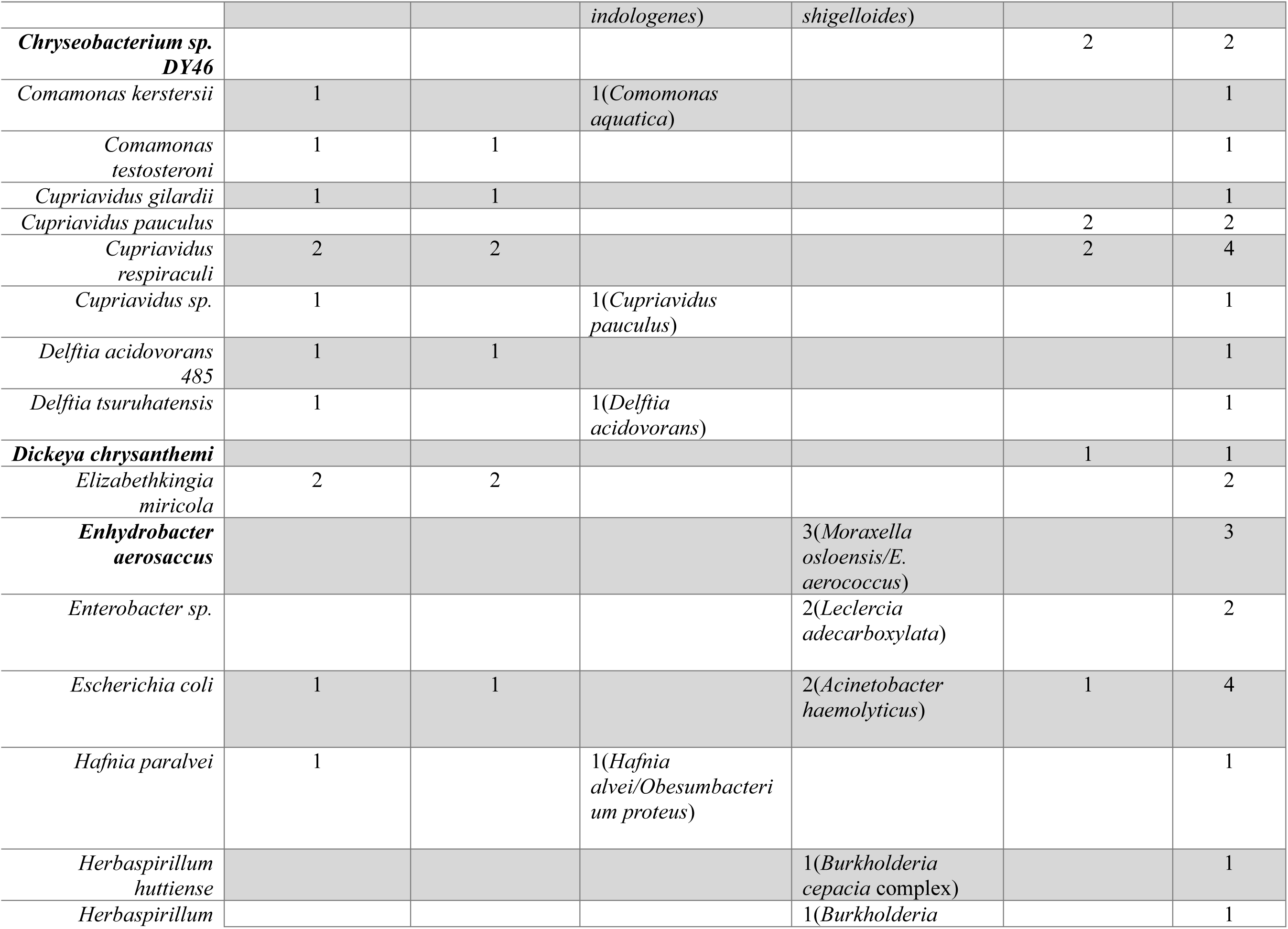
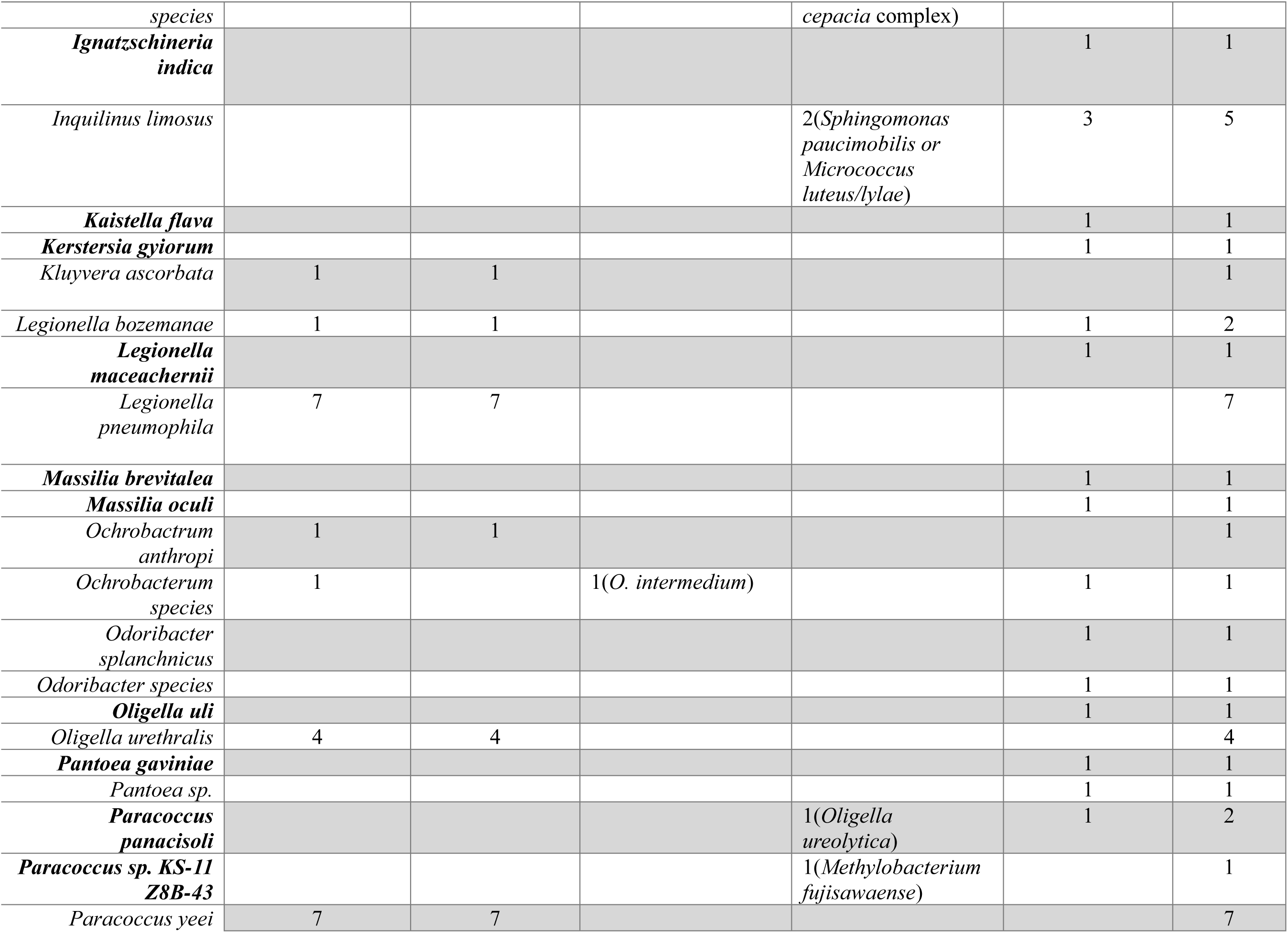
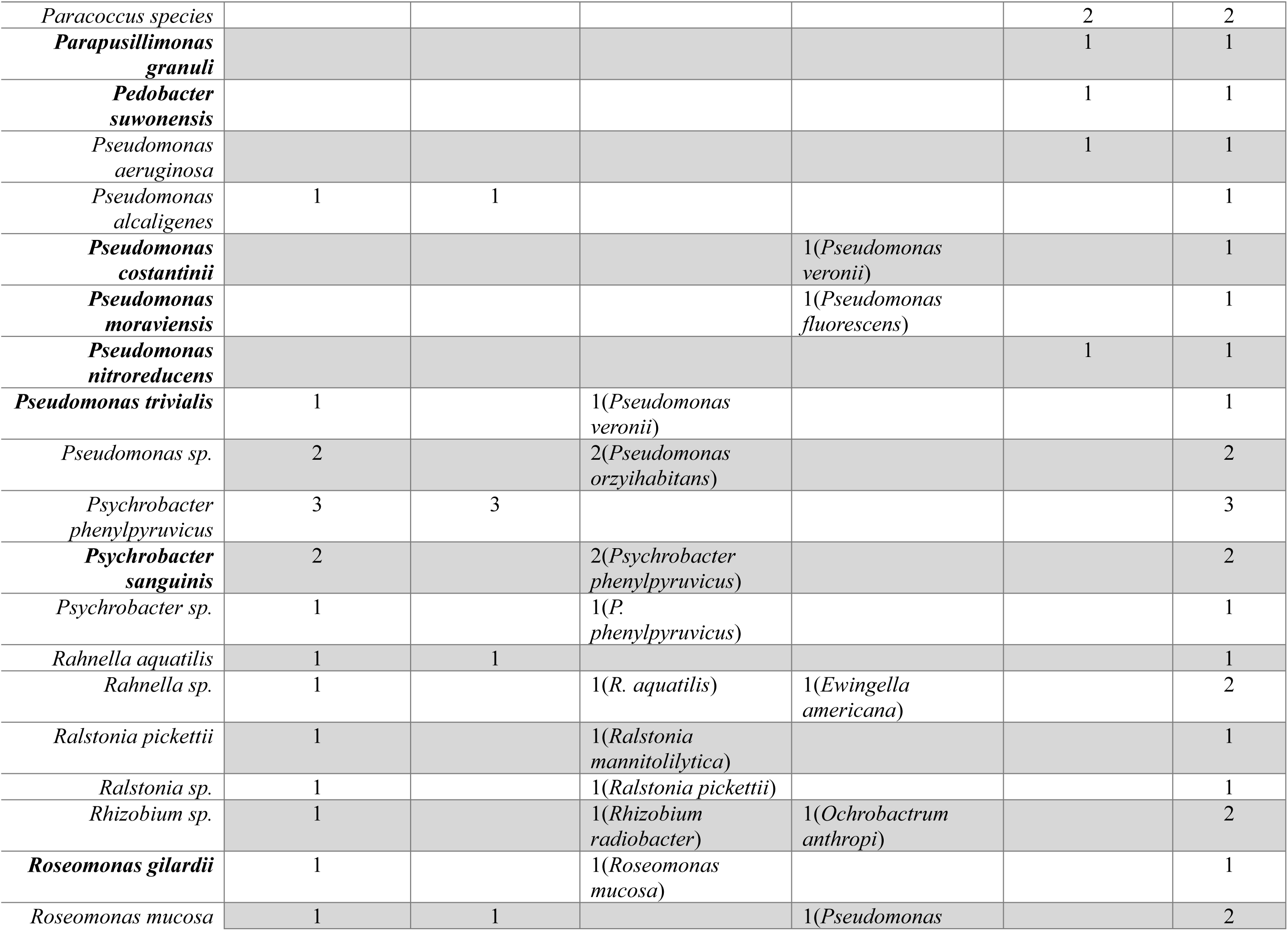
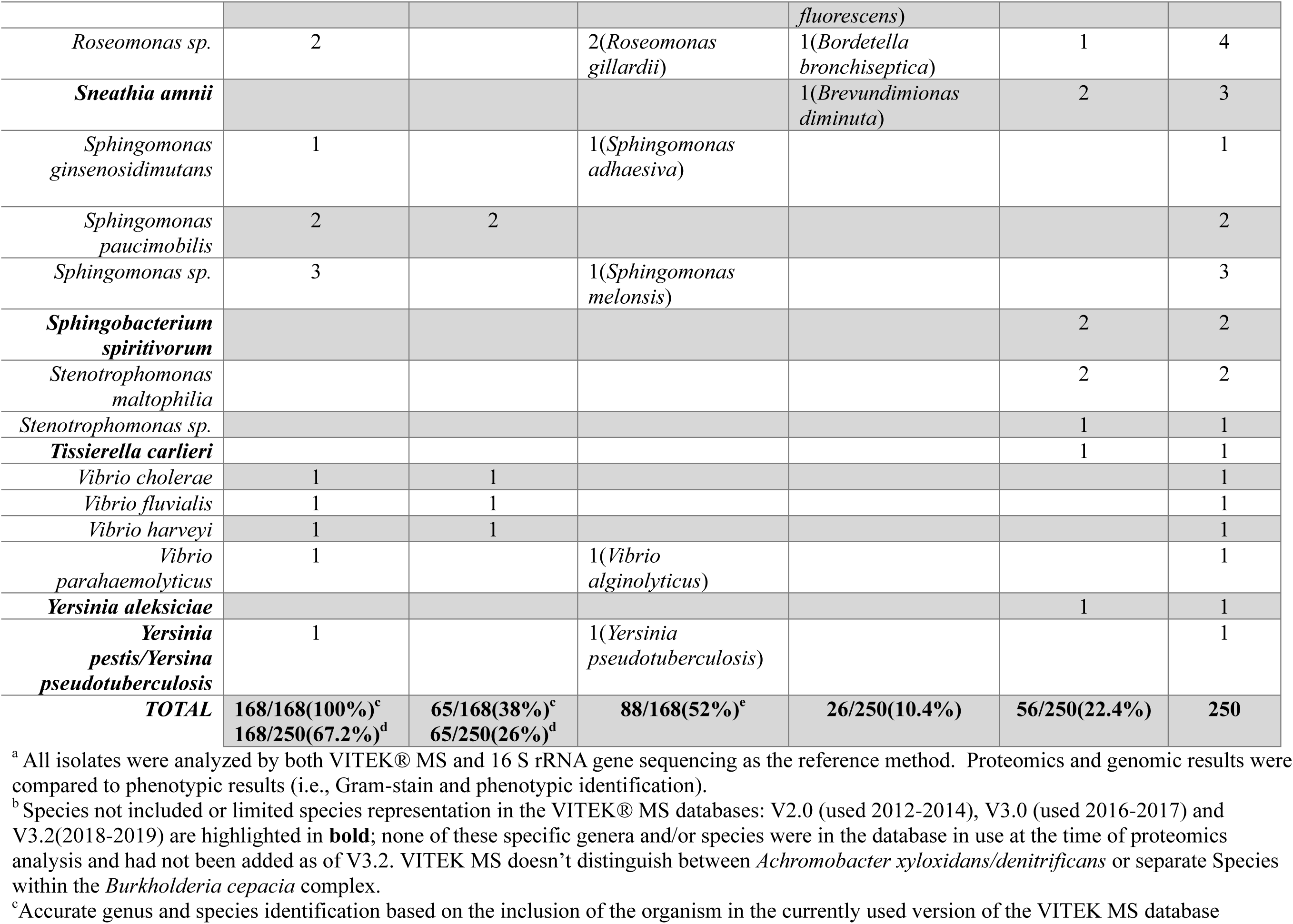
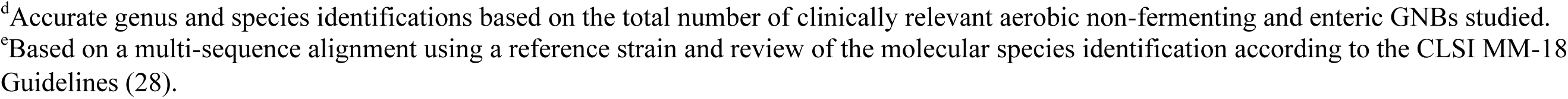
Performance of VITEK® MS compared to molecular identification of aerobic non-fermenting gram-negative bacilli (NFGNBs)

**Table 2** describes 9 genera and 25 species of fGNBs studied. Predominant genera were *Aggregatibacter* spp. (n=13, 21%), *Capnocytophaga* spp. (n=10, 16%) and *Neisseria* spp. (n=11, 18%) but a diverse spectrum of fGNBs were represented. Predominant species included *Aggregatibacter* spp. (n=16) (*A. actinomycetemcomitans*, *A. aphrophilus*, *A. kiliani* and *A. segnis*), *Capnocytophaga* spp. (n=13) (*C. canis*, *C. cynodegmi*, and *C. sputigena*) and *Neisseria* spp. (n=11) (*N. elongata*, *N. flavescens*, *N. meningitidis*, *N. mucosa*, *N. subflava* and *N. weaveri*). Accuracy pf VITEK® MS fGNB identification remained stable due to limited addition of new fGNB spectral database profiles **Table 2**.

**Table 2.**
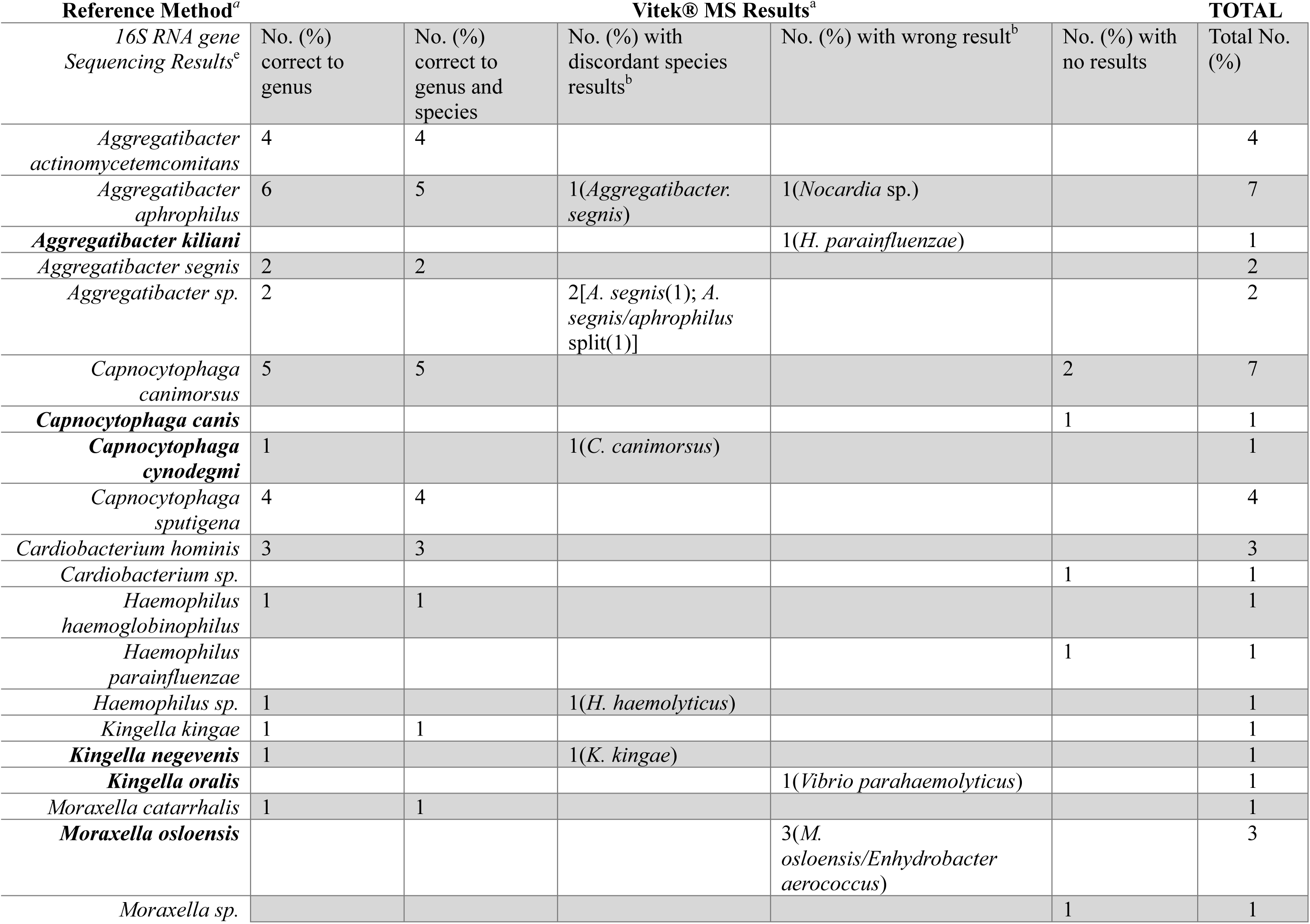
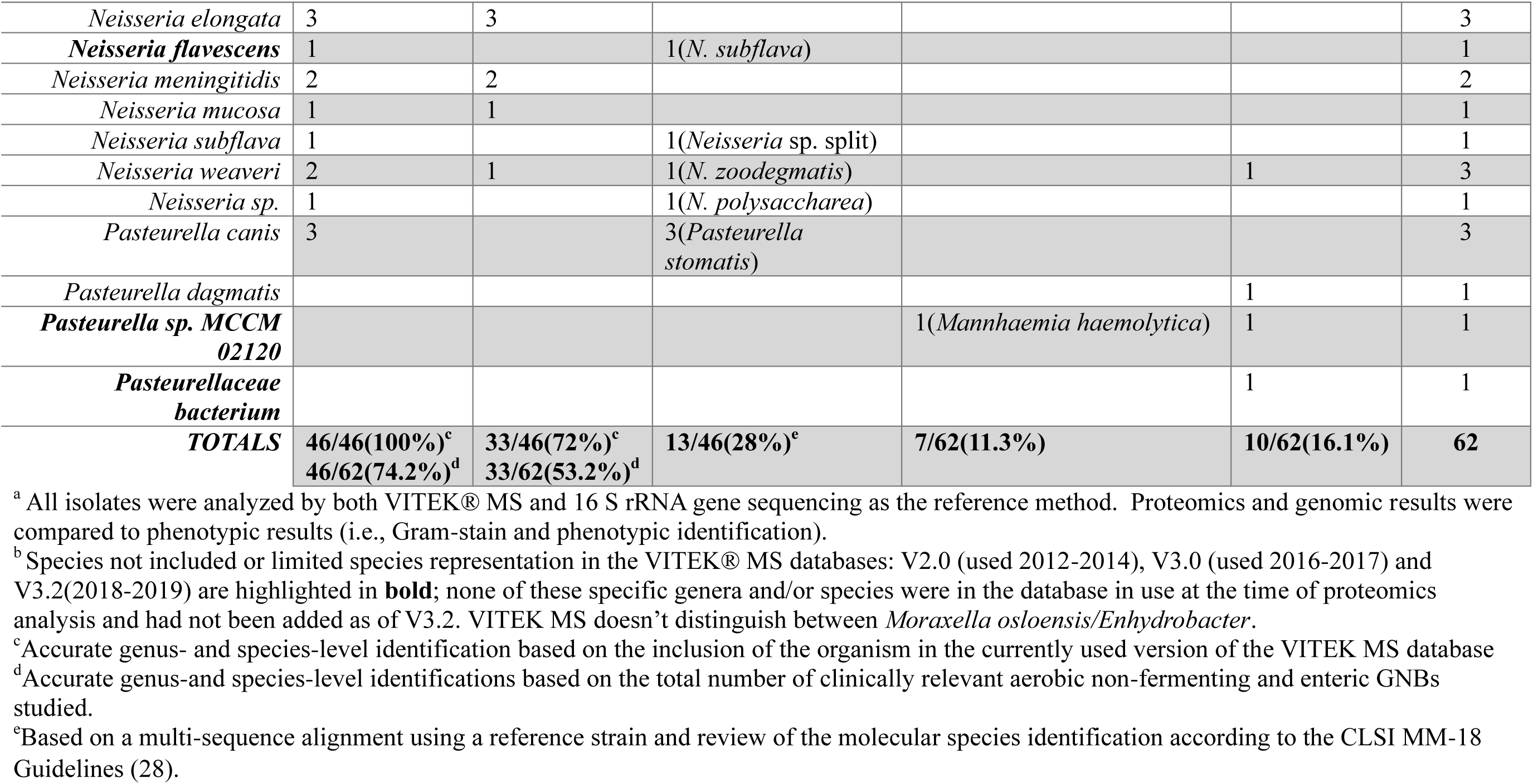
Performance of VITEK® MS for identification of fastidious gram-negative bacteria.

**Table 3** describes 2 genera (*Campylobacter* spp. (n=11, 65%) and *Helicobacter* spp. (n=6, 35%), and 11 species of CAMPB studied. Predominant species included *Campylobacter gracilis*, *C. ureolyticus* and *Helicobacter cinaedi*. Accuracy of VITEK® MS CAMPB identification improved using V 3.2 database with addition of *Campylobacter curvus* and *C. rectus*.

**Table 3.**
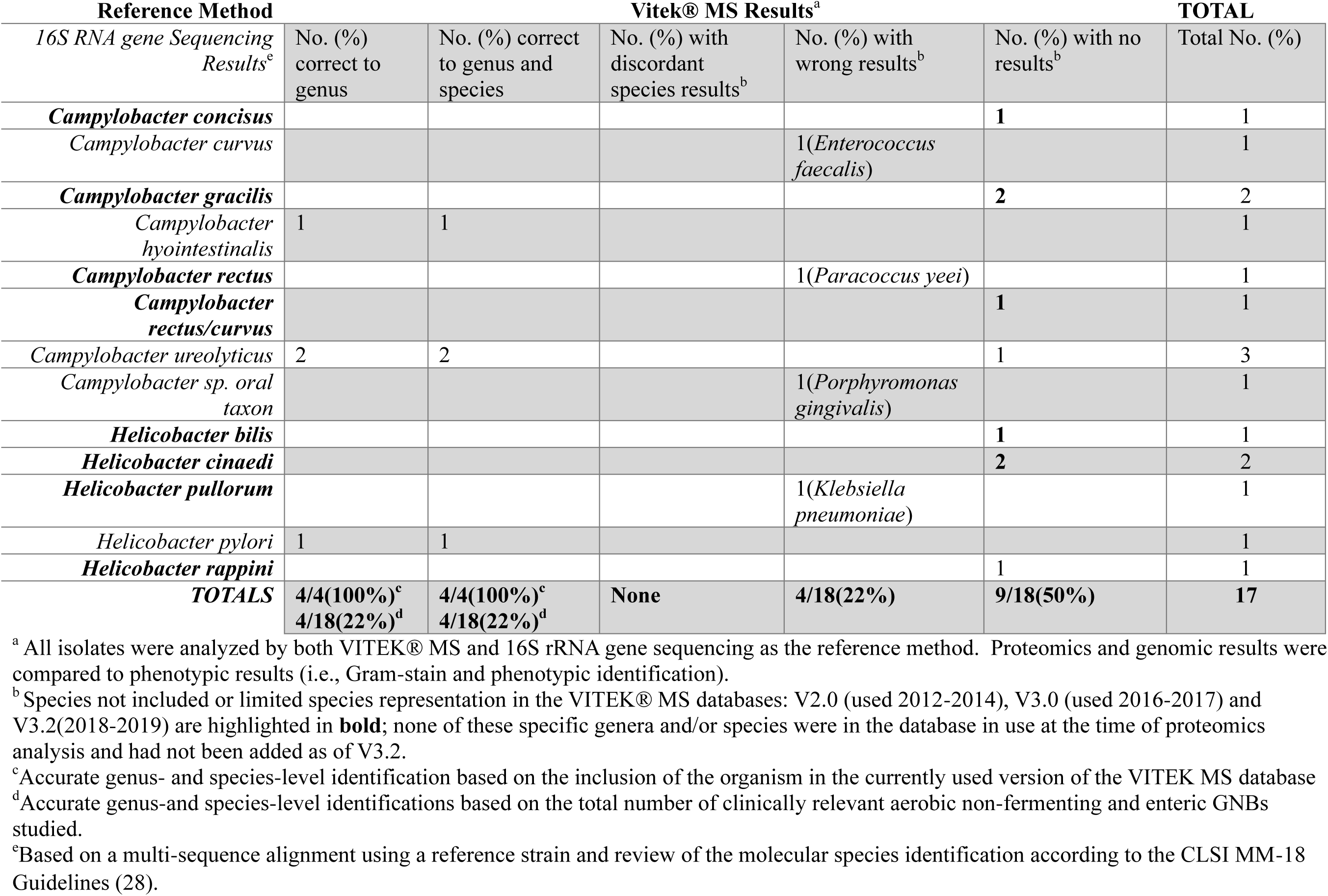
Performance of VITEK® MS for identification of Campylobacterales.

### 2.2 Performance of VITEK MS for GNB identification

Overall, VITEK MS accurately identified 218/329 (66.3%) and 102/329 (31%) of GNBs to the genus and species, respectively. Wrong identifications and no results occurred for 46(14%) and 75(22.8%) isolates, respectively. 16S accurately identified 329 (100%) and 269 (82%) to the genus and species, respectively.

**Table 1** compares performance of VITEK MS to 16S for NFGNB identification. Proteomics accurately identified 67.2% and 26% isolates to genus and species, respectively. Discordant species-level identifications are shown in **Table 1**. No result (56/250; 22.4%) or a wrong identity (26/250; 10.4%) occurred for 33% of NFGNBs because organisms were not included in VITEK MS databases (highlighted in bold in **Table 1).** Wrong identifications included *Microbacterium flavescens*, *Pleisiomonas shigelloides*, *Acinetobacter haemolyticus*, *Burkholderia cepacia* complex, *Sphingomonas paucimobilis*, *Micrococcus luteus/lylae* 50/50 split, *Oligella ureolytica*, *Methylobacterium fujisawaense*, *Ewingella americana*, *Ochrobactrum anthropi*, *Bordetella bronchiseptica* and *Brevundimonas diminuta* (**Table 1**). VITEK® MS doesn’t distinguish between *Achromobacter xyloxidans/denitrificans* 50/50 split, *Moraxella osloensis/Enhydrobacter aerococcus* 50/50 split and *Yersinia pseudotuberculosis*/*pestis*. 16S accurately identified 100% of isolates to genus and most (199/250; 80%) NFGNBS species.

**Table 2** compares performance of VITEK MS to 16S for fGNB identification. Proteomics accurately identified 74.2% and 53.2% isolates to genus and species, respectively. Discordant species-level identifications are shown in **Table 2**. No results (10/62; 16.1%) or a wrong identity (7/62; 11.3%) occurred for 26% of fGNBs because organisms were not included in VITEK MS database (highlighted in bold in **Table 2**). Wrong identifications included *Nocardia* sp., *Haemophilus parainfluenzae*, *Vibrio parahaemolyticus*, *Moraxella osloensis*, and *Mannhaemia haemolytica*. 16S accurately identified 100% of isolate to genus and most (55/62; 89%) fGNB species.

**Table 3** compares performance of VITEK MS to 16S for CAMPB identification. Proteomics accurately identified (4/18; 22%) *Campylobacterales* isolates. No discordant species-level identifications occurred. No results or wrong identity occurred for most (13/18; 72.2%) CAMPB. No results (9/18;505%) or a wrong identity(4/18; 22%) occurred for most CAMP because organisms were not included in VITEK MS database (highlighted in bold in **Table 3**). Wrong identifications included *Enterococcus faecalis*, *Paracoccus yeei*, *Porphyromonas gingivalis* and *Klebsiella pneumoniae*. 16S accurately identified 100% of isolates to genus and most (15/18; 83.3%) CAMPB species.

## 3. Discussion

This study uniquely highlights utility and pitfalls of proteomics for accurate identification of rare and unusual GNBs within a sequential clinical microbiology workflow. Although most clinically encountered GNBs were accurately identified during study period (data not shown), MALDI-TOF MS performance for study organisms was less optimal. VITEK® MS gave an accurate genus result for 66.3% of GNB isolates but gave a wrong identification or no results for 14% and 22.4% of isolate respectively. VITEK MS has difficulty identifying most *Campylobacterales* due to the limited number of *Campylobacter* and *Helicobacter* species available in the instrument’s database. Addition of *Campylobacter curvus* and *C. rectus* to the V3.2 database improved identification of these species in the later part of the study. Performance remained stable for most other types of GNBs since few organisms were added to instrument’s database during study. 16S provided an accurate genus identification for all isolates but species identification could improve with sequencing of a much larger portion of the gene up to ∼1160 base pairs; many GNBs have almost complete homology within the 16S V1-V3 gene regions (∼first 500 bp interrogated by the fast MicroSEQ™ 500 16S DNA PCR kits used)(28).

Previously reported MALDI-TOF MS verification studies showed improved proteomics performance compared to our data because of primary selection of isolates based on inclusion in an instruments’ database. Faron and colleagues performed a large multicenter evaluation of Bruker MALDI Biotyper for identification of 2,263 NGGNB isolates representing 23 genera and 61 species(29). Compared to sequencing, proteomics correctly identified 99.8% (2,258/2,263) to genus and 98.2% (2.222/2,269) to species(29). Gautam and colleagues analyzed 150 NFGNB (either *Acinetobacter baumannii, Burkholderia cepacia* complex or *Stenotrophomonas maltophilia*) isolates using MALDI Biotyper (Bruker Daltronics) following molecular confirmation and found genus and species identification agreement of 100% and 73.3%, respectively(30).

Our studies focussed on performance of MALDI-TOF MS for identification of respiratory tract NGNBs isolated from cystic fibrosis patients(7, 25, 31, 32). Plonga and colleagues assessed VITEK MS for this purpose and showed a 58.5% and 46.2% accuracy for genus compared to species identification using V2.0 to 3.1 databases(7). Accuracy improved to 89.2% and 83.6% for genus compared to species identification using instrument’s SARAMIS™ database (research use only) due to improved VITEK identification of *Burkholderia cenocepacia* and *B. contaminans*. Few reports compare proteomics for accurate identification of fGNBs. Branda and colleagues performed a multicenter validation of VITEK MS (V2.0 database) including 226 isolates representing of 89 genera and 15 species of previously sequenced fGNBs; 96% of isolates were accurately identified to genus- or species(22). Schulss and colleagues studied 150 NFGNBs and 50 fGNBs previously characterized (i.e., phenotypic methods and 16S) isolates using MALDI Biotyper, and found lower accuracy for genus (53.7%) and species (46.4%) identification despite performing direct transfer plus formic acid preparation and ethanol-formic acid extraction(33). Proteomics performance for *Campylobacterales* has not previously been reported.

Approximately one-half of proteomic errors in our study occurred because specific organisms were not included in VITEK MS databases (**Tables 1-3** highlighted in **bold**). Proteomics errors have also been reported to occur because a quality peak is difficult to distinguish with specific spectra(34). MALDI-TOF MS may also have difficulty separating some genera and species giving lower identification score related to interspecies similarities(26, 28). Many important GNBs also have closely related 16S genetic sequences and proteomics spectral profiles decreasing these methods’ ability to give an accurate identification(35). 16S gene sequencing should remain in place within our laboratory’s workflow (**Figure 1**) until sufficient proteomic spectra for rare and unusual NFGNBs, fGNBS and *Campylobacterales* are included in VITEK MS instrument databases.

Limitations of this work include enrollment of isolates from a single Canadian center albeit our laboratory performs all microbiology testing for an entire regional population. Geographic strain differences for GNBs may cause discordant identification rates, and limited overall data comparing proteomics results across MALDI-TOF MS platforms has been globally reported. Chart reviews may have improved assessment of pathogenicity of some isolates, but each case was medically assessed prior to enrollment. A caveat to clinical identification accuracy remains necessity of reporting a genus versus species; although only genus identity may suffice for patient care, rare and unusual GNB species have unique inherent or acquired multi-drug resistance profiles and epidemiologic patterns of disease.

## 4. Conclusions

Laboratories should have a workflow for identification of pathogenic unusual or rarely encountered aerobic, fastidious and *Campylobacterales* GNBs that includes 16S rRNA gene sequencing whenever proteomics cannot give a definitive identification.

## Data Availability

Data supporting the findings of this study are available from Alberta Health Services (AHS), Alberta Precision Laboratories (APL) but restrictions apply to availability of se data, were used under ethics agreement for current study, and so are not publicly available. Data are available from authors upon reasonable request and with permission of AHS/APL.

## Consent for publication

Not applicable

## Availability of data and materials

Data support findings of this study are available from Alberta Health Services (AHS), Alberta Precision Laboratories (APL) but restrictions apply to availability of se data, were used under ethics agreement for current study, and so are not publicly available. Data are available from authors upon reasonable request and with permission of AHS/APL.

## CRediT authorship contribution statement

**DL Church:** formal analysis, Conceptualization, Data curation, Methodology, Project administration. Writing – original draft, Writing – review & Editing. **T. Griener:** Methodology, Writing – review and editing. **D. Gregson:** Methodology, Writing – review & Editing.

## Acknowledgements

We would like to thank medical laboratory technologists at Alberta Precision Laboratories for their assistance with isolate testing.

